# Regulatory risk loci link disrupted androgen response to pathophysiology of Polycystic Ovary Syndrome

**DOI:** 10.1101/2025.03.26.25324630

**Authors:** Jaya Srivastava, Ivan Ovcharenko

## Abstract

A major challenge in deciphering the complex genetic landscape of Polycystic Ovary Syndrome (PCOS) lies in the limited understanding of how susceptibility loci drive molecular mechanisms across diverse phenotypes. To address this, we integrated molecular and epigenomic annotations from proposed causal cell-types and employed a deep learning (DL) framework to predict cell-type-specific regulatory effects of PCOS risk variants. Our analysis revealed that these variants affect key transcription factor (TF) binding sites, including NR4A1/2, NHLH2, FOXA1, and WT1, which regulate gonadotropin signaling, folliculogenesis, and steroidogenesis across brain and endocrine cell-types. The DL model, which showed strong concordance with reporter assay data, identified enhancer-disrupting activity in approximately 20% of risk variants. Notably, many of these variants disrupt TFs involved in androgen-mediated signaling, providing molecular insights into hyperandrogenemia in PCOS. Variants prioritized by the model were more pleiotropic and exerted stronger downregulatory effects on gene expression compared to other risk variants. Using the IRX3-FTO locus as a case study, we demonstrate how regulatory disruptions in tissues such as the fetal brain, pancreas, adipocytes, and endothelial cells may link obesity-associated mechanisms to PCOS pathogenesis via neuronal development, metabolic dysfunction, and impaired folliculogenesis. Collectively, our findings highlight the utility of integrating DL models with epigenomic data to uncover disease-relevant variants, reveal cross-tissue regulatory effects, and refine mechanistic understanding of PCOS.

## Introduction

Polycystic Ovary Syndrome (PCOS) is a multifactorial endocrine disorder characterized by abnormal LH:FSH (Luteinizing hormone: Follicle Stimulating hormone) ratios and elevated androgen levels, leading to anovulation, polycystic ovaries, and various hyperandrogenism-related comorbidities (1–3). The reproductive abnormalities in PCOS stem from disruptions in the hypothalamic-pituitary-gonadal (HPG) axis, which also contributes to other conditions like oligomenorrhoea, ovarian insufficiency, infertility, hyper- and hypogonadism, and endometriosis (4). This overlap in clinical features complicates PCOS diagnosis, prompting the establishment of multiple diagnostic criteria by the NIH, Rotterdam, and the Androgen Excess and PCOS Society. Based on a consensus, diagnosis relies on clear indications of hyperandrogenism, polycystic ovarian morphology and ovulatory dysfunction (3,5).

Decades of research on molecular mechanisms underlying the disease have identified impaired folliculogenesis and enhanced steroidogenesis in theca and granulosa cells (GCs) as key contributors to PCOS development (6). These pathways are spatiotemporally regulated by LH and FSH, secreted by the pituitary gland in response to hypothalamic Gonadotropin Release Hormone (GnRH) based stimulation (7). Moreover, PCOS often coincides with hyperinsulinemia, though the molecular origins of the association between the two is still being investigated. Hyperinsulinemia worsens hyperandrogenism by affecting adrenal androgen production and reducing sex hormone-binding globulin (SHBG) levels in the liver (8) and may also contribute to the metabolic co-morbidities such as obesity, type-II diabetes and liver dysfunction (3,9).

The genetic basis of PCOS is thought to involve impaired regulation of the HPG axis (10). Polymorphisms in genes coding for kisspeptin (Kiss1, an upstream regulator of GnRH), GnRH receptor, Anti-Mullerian Hormone (AMH), LH, FSH, and their receptors have been linked to impaired signaling in PCOS patients (11,12). However, except for *AMHR*, no functional studies directly connect these polymorphisms to the PCOS phenotype (13). Twin studies suggest an estimated 80% heritability [13], highlighting the need for detailed investigations into the molecular mechanisms underlying PCOS etiology (14).

PCOS genome-wide association studies (GWAS) across diverse populations have identified novel disease loci, including plausible candidates such as *FSHR, FSHB*, and *LHCGR*, in addition to several others with no direct association with disease phenotypes (10). Variants in the loci *of THADA, DENND1A, IRF1, FTO* etc., have significant correlations with the disease manifestation but have not been contextually studied. On the other hand, polymorphisms in reproductive hormone receptors, including androgen (AR) and estrogen receptors (*ESR1/2*), have been implicated in PCOS phenotypes (15,16); however, these associations have not been identified in GWAS studies. These observations align with the omnigenic model of complex trait regulation, where core genes directly influence the phenotype, while peripheral genes contribute through cell-type-specific regulatory networks (17). Variants exert their net impact through complex genomic and epigenomic interactions, manifesting as GWAS association signals. This highlights the need for further investigation into the regulatory networks governing phenotypic complexity of PCOS. In this context, several isolated studies offer insights into the involvement of GWAS-identified genes in PCOS pathophysiology (10). For instance, studies show that *ERBB4* and *GATA4* regulate folliculogenesis (18,19), *ZNF217* regulates androgen production in theca cells (20), and *HMGA2* promotes granulosa cell proliferation (21). Interestingly, some genes exhibit pleiotropy depending on cell-type context; *HMGA2* also regulates adipogenesis (22), and FSH influences bone density and adipose mass (4). These findings suggest that PCOS-associated variants across multiple loci impact different cell-types through hitherto unexplored molecular mechanisms, contributing to phenotypic comorbidities.

In this study, we performed a functional assessment of PCOS susceptibility loci by integrating epigenomic data, functional assays, and a deep learning (DL)-based approach to identify causal single nucleotide variants (SNVs) across eleven disease-associated cell types. We further investigated their potential influence on the molecular mechanisms underlying PCOS etiology. This approach facilitated the identification of key transcription factors (TFs) involved in folliculogenesis, androgen-mediated signaling, and ovarian development, whose binding sites are predicted to be disrupted by causal variants. Using the well-characterized regulatory locus of *IRX3*, we demonstrate how DL models combined with prior knowledge of key PCOS TFs can effectively prioritize causal variants.

## Results

### A majority of PCOS risk SNVs lie in regulatory regions and are enriched in neuroendocrine cell-types

To conduct a comprehensive analysis of the regulatory features of PCOS risk loci, we identified 91 single nucleotide variants (SNVs) from twelve GWAS studies (Table S1). This set was expanded to 1,472 SNVs, referred herein as pcosSNVs, by including variants in linkage disequilibrium (LD) with GWAS-identified variants across all superpopulations (African, American, South and East Asian, European) with an r² ≥ 0.8, obtained from SNiPA (23). Adjacent variants within 100kb were merged to define 50 genetic loci, named based on the nearest gene and/or previously associated genes in the literature (Figure 1A, Table S2). Most of these variants are located within intronic regions, with the highest density observed in the *DENND1A* and *AOPEP* loci (Figure 1A). We then assigned target genes using the ENCODE-rE2G model (https://github.com/EngreitzLab/ENCODE_rE2G) that predicts enhancer-gene interactions across various cell types by integrating enhancer activity, 3D chromatin interactions, and DNase I hypersensitivity maps. Using a threshold of 0.8 for the rE2G score predicted from the logistic regression model, we obtained 97 target genes (Table S3), many of which were not previously linked to PCOS. These genes are significantly enriched in pathways related to cell development, differentiation, and apoptosis (hypergeometric p-value 10^-6^, Figure 1B), highlighting their potential roles in the biological mechanisms underlying the five developmental stages of folliculogenesis in oocytes and granulosa cells (24).

**Figure 1:**
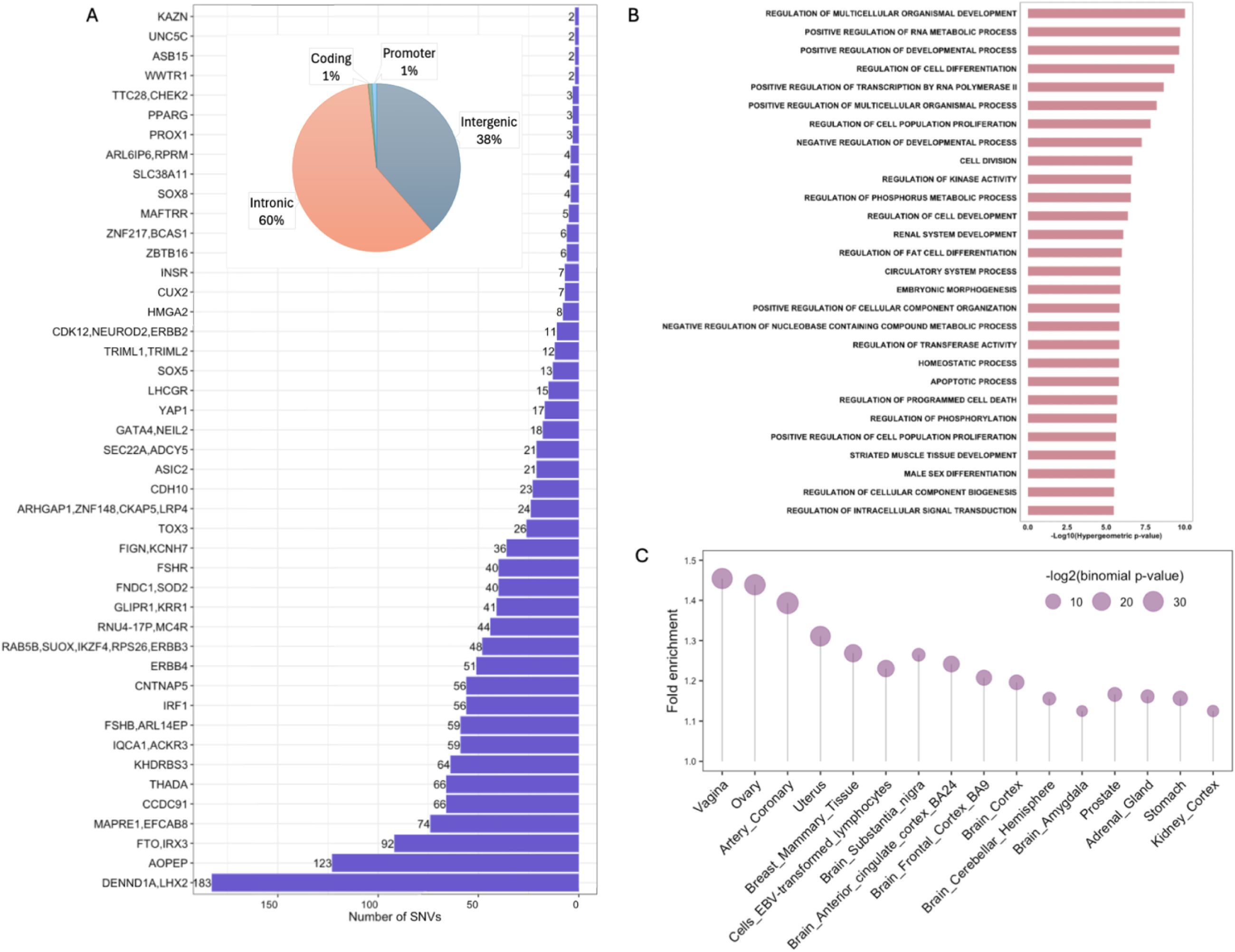
(A) PCOS susceptibility loci and their distribution in non-coding regions, (B) Gene Ontology annotations of target genes, (C) Fold enrichment of PCOS eVariants in GTEx cell-types (reported eVariants with enrichment binomial p-value < 0.01).

The diverse phenotypic comorbidities associated with PCOS typically manifest as either metabolic or reproductive abnormalities. To investigate potential differences between these subtypes, we categorized pcosSNVs into metabolic or reproductive groups based on two defined criteria: (1) trait descriptions provided in the GWAS summary statistics (Table S1), and (2) alignment of phenotypic characteristics reported in the respective GWAS with known subtype-specific features—namely, elevated AMH, LH, and SHBG levels for reproductive, and high BMI, insulin, or glucose levels for metabolic—as described in prior studies (25,26). Based on the sub-phenotype associations of GWAS susceptibility variants, we classified fifteen loci as metabolic and sixteen as reproductive (Table S4). To elucidate the regulatory mechanisms distinguishing the two subtypes, we examined the differential enrichment of TFBSs in each subtype (Figure S1A, Methods). The metabolic pcosSNVs showed significant enrichment for TFBSs of NR1D1 and *RXRB*, which regulate adipocyte differentiation (27,28), and steroid hormone nuclear receptors *RXRB*, and *THRA* (28) (binomial p-value < 0.01 vs reproductive pcosSNVs). Interestingly, binding sites of TFs involved in the development and differentiation of GCs during folliculogenesis, such as *CPEB1* and *FOXL2*, are enriched in the metabolic subtype. This suggests their role in also driving metabolic abnormalities associated with PCOS, aligning with previous observations where CPEB1 has also been shown to associate with obesity (29). Conversely, the reproductive subtype is enriched for TFBSs of *NR5A1*, and *NR1H4* (binomial p-value < 0.01 vs metabolic pcosSNVs), all of which have well-documented roles in steroidogenesis and folliculogenesis (30,31). Furthermore, the two subtypes are enriched for variants from distinct GWAS trait groups, correlating with their phenotypic traits (Figure S1B, Methods). For example, reproductive subtype variants exhibit significant enrichment for endocrine traits like endometriosis and uterine fibroids (binomial p-value < 10^-15^ vs pcosSNVs), while metabolic subtype variants show significant enrichment for obesity, BMI, and cholesterol (>5-fold, binomial p-value < 10^-30^ vs pcosSNVs).

We next investigated the functional impact of pcosSNVs through their association with changes in gene expression characterized by the GTEx consortium (32). Among 1,472 pcosSNVs, 832 overlapped with cis-eQTLs, termed eVariants (Table S5). Two-thirds of these pcosSNVs are shared across multiple cell-types (we use the term cell-types herein synonymously with tissues defined in GTEx), meaning they influence gene expression in multiple cell types. In contrast, the remaining variants, such as those in the *FSHR* and *ERBB4* loci, are cell-type specific (Figure S2a). The number of target genes scaled almost linearly with the number of affected cell types (Spearman correlation: 0.71, Figure S2b), suggesting that shared eQTLs may contribute to distinct cell-type specific regulatory networks by regulating different genes in different cell-types. Notably, eVariants in the locus of the GATA4 gene were linked to 39 genes across 49 cell types (Figure S2b, Table S6). 4% of eVariants targeted long non-coding RNAs (Table S5), suggesting their role as potential trans-eQTLs (33). Meanwhile, the remaining 640 pcosSNVs that were not identified as GTEx eVariants belong to 18 susceptibility loci, including that of *CNTNAP5*, *ASIC2*, and *CDH1*, (Table S6), likely due to low target gene expression or restricted function in specific cell states or developmental stages in the dynamic transcription landscape that not captured in bulk tissue analysis (32,34). These pcosSNVs may play key spatiotemporal roles in mediating GnRH response in the hypothalamus, pituitary regulation, and follicular phase progression in PCOS (32).

We also examined the enrichment of PCOS eVariants across GTEx cell types. Compared to a randomly selected set of 832 eQTLs (excluding PCOS eVariants), PCOS eVariants were found to be enriched in brain cell types, reproductive hormone-producing tissues such as the ovary and adrenal gland, as well as hormonally influenced tissues like the breast and prostate (binomial p-value < 0.001, Figure 1C). Since many PCOS eVariants are shared across cell-types, they likely influence regulatory networks by mediating interactions between cell-type-specific and ubiquitous transcription factors (TFs), thereby invoking cell-type-specific regulatory pathways that contribute to distinct phenotypic outcomes in different cellular contexts (35). For example, *PPARG*, a susceptibility locus, plays a central role in regulating lipid metabolism, adipocyte differentiation, gluconeogenesis, folliculogenesis, and steroidogenesis through multiple TFs that are critical regulators of these biological processes (www.kegg.jp/pathway/map=map03320) (36,37). This suggests that causal variants within this locus may contribute to distinct phenotypic outcomes through pleiotropic effects across multiple cell types. Given these complexities, investigating disease-causal variants and their cell-type-specific effects on downstream signaling pathways may help elucidate the mechanisms underlying the diverse phenotypic manifestations of PCOS.

### A deep learning model for prioritizing disease causal variants across causal cell-types

The challenge of characterizing the cell-type-specific impact of thousands of susceptibility variants in complex traits and diseases has led to the development of computational approaches for inferring causal variants. DL models have been particularly effective in predicting variant effects on gene regulation by integrating diverse cell-type-specific epigenomic features (38,39). We previously developed a convolutional neural network based DL model, TREDNet, which can predict the effects of non-coding variants on enhancer activity (40). This two-phase DL model was demonstrated as a successful approach in prioritizing causal variants of type 2 diabetes and autism (40,41). Building on its success, we applied TREDNet to investigate the regulatory mechanisms underlying PCOS.

We adapted TREDNet to predict allele-specific enhancer activity of PCOS-associated SNVs (pcosSNVs) across causal cell types implicated in PCOS. Based on the biological origins of folliculogenesis, androgenesis, and signaling pathways involving SHBG, insulin signaling, and adipogenesis that are known to be disrupted PCOS, we identified eleven causal cell types for analysis with available epigenomic profiles: KGN (used as a proxy for granulosa cells due to limited epigenomic data from primary granulosa cells), ovary, fetal brain, adrenal gland, pancreas, adipocytes, liver, brain microvascular endothelial cells (BMEC), mammary epithelial cells, and human umbilical vein endothelial cells (HUVEC). In the absence of epigenomic profiles from the pituitary and hypothalamus, we included fetal brain, and selected BMEC, mammary epithelial cells, and HUVEC as granulosa cell proxies based on their epithelial or endothelial characteristics and similar H3K27ac profiles (Jaccard similarity index, Table S7) (42). Additionally, we incorporated WTC11, a developmental cell line, to capture causal variants active during early development, as fetal development has been implicated in PCOS onset later in life (43). We trained our model using cell-type-specific putative enhancers identified by co-occurrence of the H3K27ac histone modification with DNase hypersensitive sites (DHS), as proxies for active enhancers (Table S8, Methods). The DL models demonstrated robust performance, achieving an area under the receiver operating characteristic curve (auROC) ranging from 0.9 to 0.98 and an area under the precision-recall curve (auPRC) ranging from 0.54 to 0.84 across the eleven cell types (Figure 2A).

**Figure 2:**
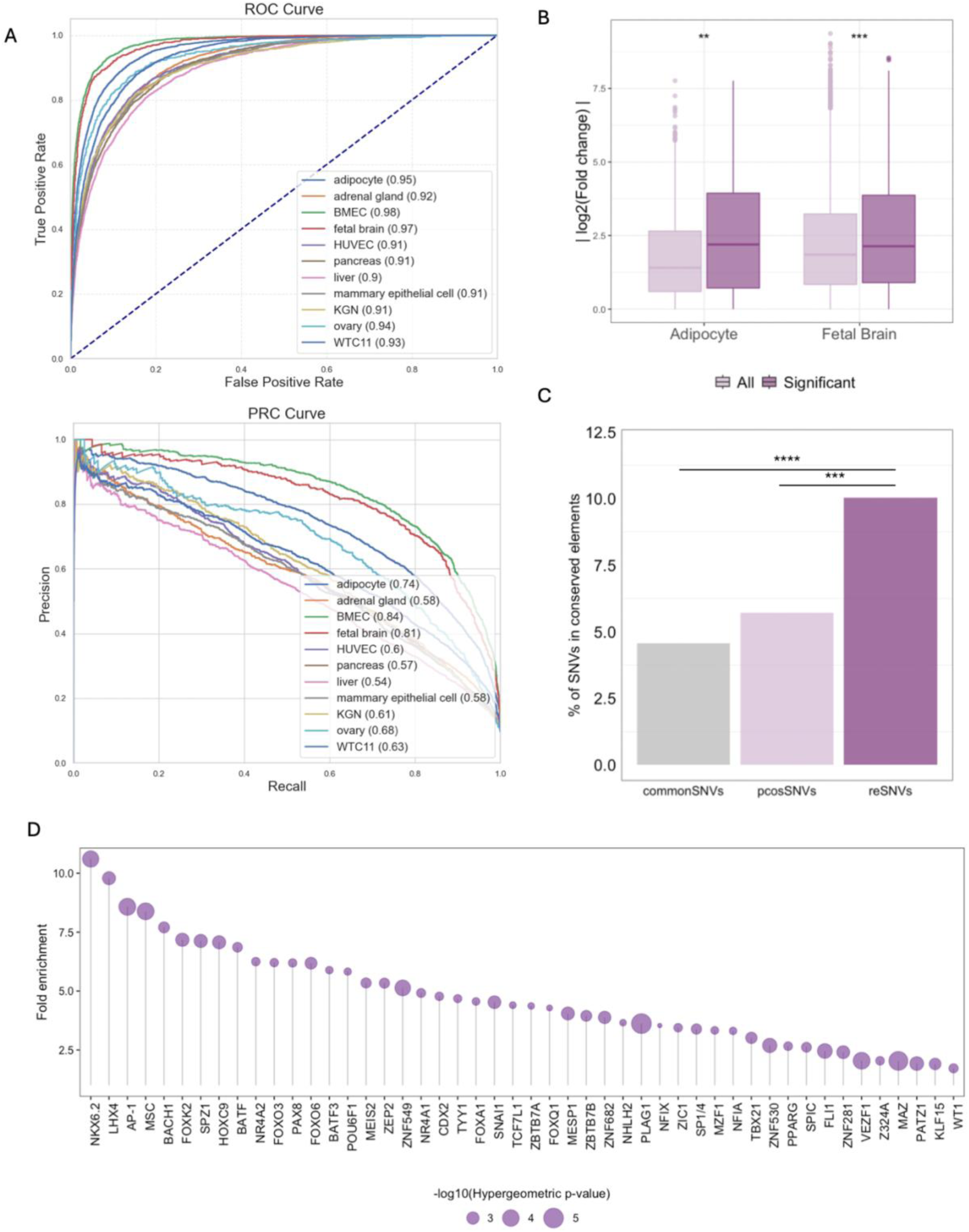
(A) ROC and PRC curves of eleven cell-type specific TREDNet models, (B) A comparison of fold change (alternate / reference allele) in TREDNet scores between all variants and those exhibiting significant change in enhancer activity in MPRA, using Wilcoxon test (C) Fraction of SNVs overlapping with phastCons elements conserved across 30 primates, (D) TFs enriched among reSNVs compared with control SNVs (hypergeometric p-value < 0.01). (ns: p > 0.05, *: p <= 0.05,**: p <= 0.01,***: p <= 0.001)

To evaluate TREDNet’s ability to identify causal variants, we examined the correlation between TREDNet-predicted differences in allele-specific enhancer activity and those determined through a massively parallel reporter assay (MPRA) in the developing human brain and stem cell-derived adipocytes using our model trained on fetal brain and adipocytes (44,45). We compared allele-specific TREDNet scores across all assayed alleles and those showing significant changes in reporter activity (Methods) and observed a significantly higher fold change in TREDNet scores for the latter group (Mann-Whitney test p-value = 0.001 for adipocytes and 10^-5^ for fetal brain, Figure 2B). These findings highlight TREDNet’s robustness in predicting causal variants across different cell types.

Next, we evaluated the impact of pcosSNVs within active regulatory regions across the eleven selected cell-types, scoring both reference and risk alleles in all cell-type-specific models (Methods). For pcosSNVs located in active regulatory regions marked by H3K4me1, H3K27ac, or DNase/ATAC-Seq, we classified strengthening alleles as those with scores below the threshold (determined at 10% FDR) for the reference allele and above for the risk allele, while damaging alleles followed the opposite criterion. Applying this approach, we identified 309 pcosSNVs with predicted allelic differences in activity, termed reSNVs (Table S9). These reSNVs were significantly enriched in conserved elements compared to both pcosSNVs and 13 million common SNVs from the 1000 Genomes catalog (binomial p-value = 0.0002 and 10^-9^, respectively, Figure 2C).

To assess the regulatory impact of reSNVs, we examined the enrichment of TFBSs. The regulatory effect of a TF was quantified by comparing the density of its binding motifs overlapping with reSNVs to a background set of control SNVs. Specifically, we quantified the abundance of transcription factor binding sites (TFBSs) affected by these variants and compared it to a control set of TFBSs corresponding to 71,000 SNVs located within 100 kb of pcosSNVs (Methods). This localized background enabled us to investigate the regulation of target genes within the context of PCOS-specific biological processes, particularly for ubiquitously expressed genes. Several TFs showed significant enrichment at reSNV loci, including *FOXA1*, a pioneer factor in estrogen and androgen signaling (46); *LHX4*, involved in pituitary development (47); *NHLH2*, associated with GnRH signaling (48); *WT1*, a regulator of granulosa cell proliferation (49); PLAG1, involved in oocyte reserve maintenance (50); and *NR4A1*, which regulates steroidogenesis (51) (hypergeometric p-value < 10^-2^, Figure 2D). Notably, we observed a 2.6 fold enrichment of PPARG binding sites, a significant finding given PPARG’s role as a known susceptibility locus for PCOS. We also found enrichment of TFs associated with neuronal signaling, such as TBX21, POU6F1, and NKX6.2. While not previously linked to PCOS, these TFs represent promising candidates for involvement in neuroendocrine regulation. These findings highlight the capacity of our model to identify transcriptional regulators with potential functional roles in the diverse phenotypic manifestations of PCOS.

Our deep learning-based approach identified 20% of the pcosSNVs as potential regulatory variants, effectively narrowing down causal variants in loci such as *ERBB4*, *LHCGR, MC4R*, etc (Figure S3). For example, among 51 variants in the *ERBB4* locus, we identified rs79230362 as an enhancer-disrupting variant in HUVEC cells (Table S9). This variant is in LD with the GWAS SNP rs113168128 and is predicted to disrupt the binding site of the *ELK1:SREBF2* motif complex (Figure S4). Given the established role of *SREBF2* in steroidogenesis (52) and the highly cell-type-specific expression of ELK1 in granulosa cells (Figure S4), this variant likely affects *ERBB4* expression, a key regulator of the oocyte microenvironment during folliculogenesis (18). Similarly, in the *MC4R* locus, we identified rs17773430 as a causal enhancer-disrupting variant in WTC11 cells (Table S9). *MC4R* is a critical component of the melanocortin pathway and a well-established obesity susceptibility gene that is also linked to PCOS. Knockout studies of *MC4R* in mice result in both obesity and infertility phenotypes, highlighting shared regulatory architectures underlying these conditions (53). rs17773430 is predicted to disrupt the binding site of *TBX2/TBXT*, TFs responsible for the development of hypothalamus-pituitary axis (30). Given that reduced *MC4R* levels are associated with lower LH levels (54), this variant likely contributes to PCOS etiology through its impact on HPG axis.

On the other hand, multiple reSNVs were identified in the locus of *DENND1A*, *FTO* and *MAPRE1* (Figure S5). The significant overlap of reSNVs in *DENND1A* and *MAPRE1* locus with active regulatory regions in the fetal brain and WTC11 suggests their potential role in disease manifestation during early development. Notably, a reSNV in the *MAPRE1* locus, rs187178, was validated as an enhancer-disrupting variant in the fetal brain and functions as an eQTL for the neighboring gene *DNMT3B*, which regulates dynamic methylation transitions during folliculogenesis (24). In total, we identified 12 reSNVs that have been experimentally validated as enhancer disrupting variants in adipocytes and fetal brain through MPRA studies (Table S10) (44,45).

Of note, epigenomic data from fetal brain used by the DL model failed to capture the regulatory impact of pathogenic variants in the *FSHB* locus, including rs10835638 and rs11031006, which have been experimentally shown to reduce *FSHB* expression restricted to the pituitary gland (55). This underscores the necessity of incorporating additional, relevant cell types for a more comprehensive study of the regulatory landscape of PCOS, when experimental characterization of chromatin marks becomes available for these cell types.

### reSNVs are more likely to exert pleiotropic effects across multiple cell types by downregulating the expression of their target genes

We further explored the functional impact of reSNVs by examining their association with gene expression using eQTL data from GTEx. Among reSNVs that also act as eVariants, hereafter referred to as reVariants, we observed significantly greater enrichment in the brain, liver, adrenal gland, and pancreas compared to pcosSNVs (Figure 3A), implicating these tissues as key cell types affected by reSNVs. The proportion of causal cell types impacted by reVariants was significantly higher than that impacted by otherSNVs (i.e., SNVs not prioritized by TREDNet in causal cell types) (Figure 3B, Mann-Whitney p = 5.78×10⁻³). Within these enriched cell types, reVariants were associated with significantly stronger downregulation of gene expression relative to otherSNVs, as measured by normalized effect sizes from GTEx (Figure 3C, Mann-Whitney p = 1.88×10⁻⁹). In contrast, no significant difference was observed in gene upregulation effects (Figure 3C, Mann-Whitney p = 0.91). These findings suggest that reSNVs primarily exert their regulatory effects through downregulation of gene expression.

**Figure 3:**
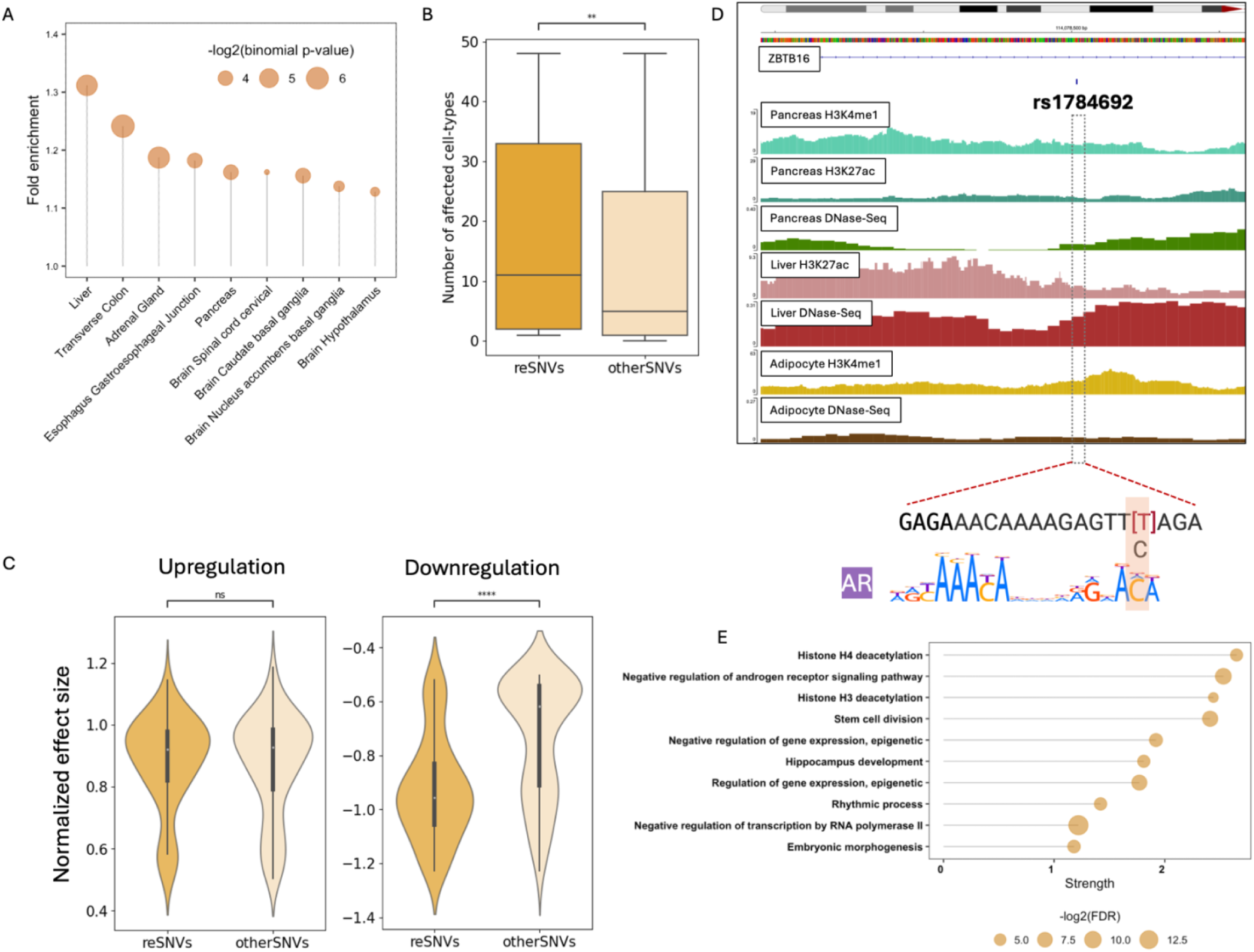
Regulatory impact of reSNVs prioritized by TREDNet. (A) Fold enrichment of reSNVs compared to pcosSNVs across cell-types (binomial p < 0.01). (B) Comparison of the number of GTEx cell-types impacted by reSNVs versus otherSNVs, (C) Normalized effect size of reSNVs versus otherSNVs. Left and right panels show differences for downregulating (NES ≤ –0.5) and upregulating (NES ≥ 0.5) variants, respectively. (D) Genomic overlap of an intronic reSNV (rs1784692) at the ZBTB16 locus with epigenomic features from cell types where it exhibits predicted allele-specific activity. The affected Androgen Receptor (AR) motif is shown below. (E) Functional enrichment of biological processes in the ZBTB16 protein interaction network (STRING database). The plot shows the top 10 terms (FDR < 0.001), with enrichment strength calculated as log₁₀(observed/expected). (ns: p > 0.05, *: p <= 0.05,**: p <= 0.01,***: p <= 0.001)

The most significant downregulatory effect was observed at the RAB5B–SUOX–RPS26 locus, where reSNVs were linked to reduced expression of RPS26 in multiple cell-types including the ovary, hypothalamus, and liver. RPS26 is a ubiquitously expressed ribosomal protein whose downregulation in the ovaries impairs oocyte growth and premature ovarian failure (56), a hallmark of PCOS. Notably, one reVariant in this region, rs3741499, which shows a large negative effect size on RPS26 expression (Figure S6), is predicted to disrupt binding of PROX1 (Table S9), a PCOS risk gene involved in lymphatic vessel formation around oocytes (57), suggesting a plausible mechanism for impaired oocyte maturation.

We further investigated pleiotropy by examining the ZBTB16 locus. Although no eQTLs overlap with variants in this locus, they were predicted to exert strong differential enhancer activity across multiple cell types (Table S9). Notably, rs1784692, located in an intron of ZBTB16, demonstrated the highest predicted enhancer-strengthening effect in the pancreas, adipocytes, WTC11, and liver (Figure 3D, Table S9). The T→C polymorphism enhances *AR* receptor binding, suggesting a possible association of this locus with cell-type-specific androgen response functions, such as insulin secretion in the pancreas (58), and regulation of adipocyte differentiation (59). While ZBTB16 has not been previously implicated in PCOS, its protein interaction network is enriched for components of androgen signaling (Figure 3E). These observations suggest that ZBTB16 may act as a susceptibility locus involved in androgen-mediated regulatory pathways disrupted in PCOS.

In conclusion, reSNVs prioritized by TREDNet offer valuable insights into disease-associated regulatory mechanisms and highlight the potential role of risk genes hitherto uncharacterized in PCOS etiology.

### The FTO locus demonstrates disruption of an androgen mediated network pleiotropy

The regulatory locus within the intronic region of *FTO* is a well-known susceptibility locus with significant implications in obesity and diabetes. Notably, it has been experimentally validated to function as a distal enhancer of *IRX3*, a TF in PCOS-associated susceptibility loci (60,61). We hypothesized that this locus may have broader pleiotropic effects across different cell types due to variations in the expression of *IRX3*, which may influence multiple biological pathways (62). Interestingly, the PCOS susceptibility variants localize in the genomic region regulating *IRX3* (chr16:53731249–54975288) (63), suggesting that *IRX3* is likely the target gene of the PCOS susceptibility locus as well (Figure S7).

We identified 12 reSNVs exhibiting significant fold changes across nine cell types (Figure S8). Among these, three variants— rs1421085, rs9940646 and rs9940128—have been validated by MPRA studies to show allelic changes in enhancer activity in mouse preadipocyte and neuronal cell lines (61), further supporting the predictive accuracy of TREDNet in identifying causal variants. Interestingly, we predicted that rs1421085 additionally upregulates enhancer activity in BMEC by potentially modulating the binding site of *ONECUT2* (Figure 4A), a suppressor of androgen receptor signaling which was recently identified as a marker of follicle growth (64,65).

**Figure 4:**
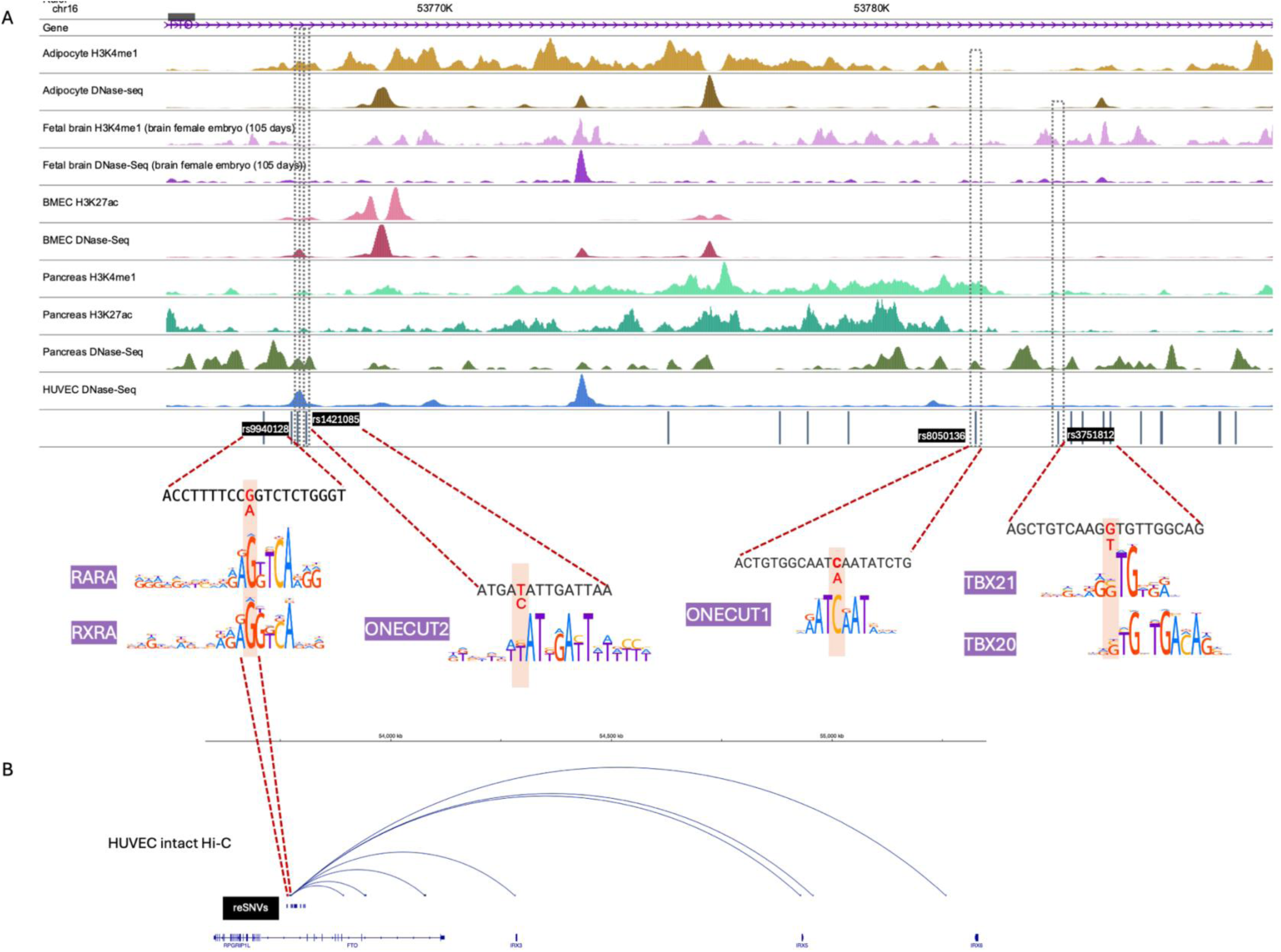
reSNVs in FTO locus exhibiting significant fold change in TREDNet predicted enhancer activity. (A) Overlap of reSNVs with active regulatory regions of pathogenic cell-types (B) Intact Hi-C map of chromatin interactions from reSNVs in FTO locus in HUVEC (doi:10.17989/ENCSR788FBI)

Additionally, we identified another variant within the same locus, rs8050136, which is predicted as a causal variant in the pancreas and liver (Figure 4A, S8). This variant functions as an eQTL for *IRX3* in the pancreas, where IRX3 regulates the conversion of beta to epsilon cells, directly linking it to type 2 diabetes (66). Notably, rs8050136 is also predicted to disrupt the binding site for *ONECUT1*, a transcription factor critical for pancreatic development (Figure 4A). Together, these findings suggest that rs8050136 may serve as another causal variant for type 2 diabetes, possibly preferentially in PCOS patients.

To address the association of these variants with PCOS, we focused on a previous study that identified *IRX3* and another gene in this susceptibility locus, *IRX5*, as key regulators of folliculogenesis in granulosa cells (67). Using evidence from granulosa like cells, BMEC and HUVEC, we hypothesize that variants in this locus lead to impaired folliculogenesis, consequently disrupting androgen production in the causal cell type—likely granulosa cells—through the dysregulated action of *IRX3/IRX5*. This disruption in androgen production may have pleiotropic effects on other cell types where these genes function within the androgen-responsive network. In this regard, rs9940128 emerges as a plausible causal variant as it forms chromatin contacts with promoters of *IRX3* and *IRX5* (Figure 4B) and is predicted to cause a significant fold change in enhancer activity in BMEC and HUVEC (Figure S8). Furthermore, the allelic effects of variants in this locus may also impact *IRX3/5*-mediated functions in hypothalamic neurons (Figure S7), as demonstrated in mice (61). To explore this further, we analyzed the impact of these variants in fetal brain and found that rs3751812 is located within binding sites of T-box family TFs (Figure 4A). Members of the T-box family play a critical role in the commitment of hypothalamus and pituitary lineages from neuronal precursors (30,68). However, given the short temporal window of expression of these TFs in neuronal development, inferring causal mechanisms remains challenging. This highlights the necessity of using epigenomic datasets across different developmental timepoints for a comprehensive investigation.

## Discussion

Our limited understanding of the regulatory landscape of PCOS stems from its complex genetic architecture, which presents with heterogeneous phenotypes across different cell types, individuals, and populations. This complexity has necessitated evolving diagnostic criteria as our knowledge of the underlying pathophysiology expands. Several key questions remain unresolved, including the genetic and molecular origins of reproductive and metabolic dysfunction, the role of androgens and other hormones in regulatory pathways, and the inheritance patterns affecting both males and females. To date, GWAS have identified 50 genomic loci associated with PCOS across diverse populations (Table S2). While the functional significance of genes such as *ERBB4*, *PPARG*, and *IRX3* has been well established, leading to the use of their agonists as potential treatments (18,37,67), the precise molecular mechanisms remain elusive. Additionally, advancements in whole-genome and exome sequencing continue to uncover novel loci, further complicating our understanding of PCOS and highlighting the need for a deeper exploration of the core regulatory mechanisms driving its pathophysiology.

Leveraging extensive genetic and epigenetic data, we sought to identify key mechanisms linking PCOS susceptibility loci to disease etiology. We found that reSNVs prioritized by our model are significantly enriched for TFBSs associated with folliculogenesis, including those of WT1, NHLH2, and FOXA1. Notably, reSNVs also show enrichment for the binding sites of PROX1 and PPARG, both of which are also PCOS risk genes. These findings underscore the importance of dissecting the underlying gene regulatory networks, where disruptions at specific nodes (genes) or edges (regulatory interactions) may give rise to a spectrum of molecular outcomes that contribute to the heterogeneity of PCOS severity and phenotypic presentation. Our results also highlight the need for further characterization of TFs, especially those involved in neuronal signaling, such as *TBX21, LHX4*, etc., along with their interactions with hormonal receptors, to gain deeper insights into cis- and trans-regulatory mechanisms disrupted in PCOS pathophysiology.

The established role of the HPG axis (69) in regulating circulating reproductive hormone levels highlights the hypothalamus, pituitary, adrenal gland, and ovarian granulosa and theca cells as key mediators of PCOS pathophysiology. However, PCOS manifestations extend beyond the neuroendocrine system, impacting peripheral tissues such as the pancreas, adipocytes, liver, and heart. This suggests that dysregulation of hormonal signaling, particularly androgens, may have widespread effects through both direct and pleiotropic mechanisms. Given the broad expression of the androgen receptor, disruptions in androgen signaling may contribute to metabolic dysfunctions—such as insulin resistance and altered adipogenesis—independent of classical reproductive symptoms like oligomenorrhea. Our findings support this expanded framework and reveal potential mechanisms by which altered androgen signaling leads to systemic effects. Accordingly, we propose two categories of pathogenic cell types: (a) primary cell types, involved directly in steroidogenesis, folliculogenesis, and reproductive hormone biosynthesis; and (b) secondary cell types, which are affected by the pleiotropic activity of risk variants or by downstream hormonal dysregulation (Figure S9). By prioritizing variants that disrupt PCOS relevant TF binding sites at susceptibility loci, we highlight the importance of TFs interacting with hormone receptors—particularly androgens—as key modulators of PCOS-related dysfunction.

The identification of multiple reSNVs at several susceptibility loci is suggestive of regulatory mechanisms wherein one-gene can be regulated by multiple enhancers, according to which, the expression of a target gene can be influenced by more than one variant (61,70). For example, two distinct variants in the *FSHB* locus, rs10835638 and rs11031006, alter *FSHB* expression, ultimately contributing to infertility (55). These variants may occur in different individuals, leading to distinct, individual-specific phenotypes depending on the cell-type-specific networks they modulate in a pleiotropic manner. In addition, the potential pleiotropic impact of disease-associated variants in non-pathogenic cell types is often buffered by robust regulatory networks, preventing overt disease manifestation. This suggests that assessing polygenic risk scores may be necessary to fully understand their contribution to disease susceptibility. Given that variants in the *FTO* locus have high minor allele frequencies (>0.4), which far exceed the prevalence of PCOS, it is evident that the disease phenotypes emerge from the cumulative effects of multiple dysregulated genes and pathways. Further investigations into polygenic interactions and gene-environment influences will be essential to expand our understanding of the complexity of PCOS.

The susceptibility loci of PCOS implicate genes such as *ZBTB16*, *AOPEP*, *THADA*, and *CCDC91* (Figure 1A), which are ubiquitously expressed, raising the question of how disease-specific variants selectively affect certain cell types. At the molecular level, follicle progression involves signaling pathways like TGFβ, Hippo, Wnt, and mTOR, which regulate fundamental processes such as cell proliferation, differentiation, and apoptosis (7). Why, then, do complex diseases manifest in only a subset of susceptible cell types? In the case of *ZBTB16*, we predicted that rs1784692 strengthens enhancer activity by increasing the binding affinity of *AR*, thereby implicating *ZBTB16* in downstream pathways of androgen signaling. This suggests that perturbations in disease-relevant TF interactions, specific to causal cell types, disrupt molecular networks in a way that surpasses compensatory mechanisms in other cell types, thereby making certain cells uniquely vulnerable. Consequently, transcription factors act as primary responders to disease-associated alterations, preceding the genes they regulate, and may therefore serve as more informative markers of disease susceptibility than the genes themselves.

Our analysis of the PCOS regulatory landscape reveals unifying molecular mechanisms underlying disease phenotypes. However, a more comprehensive understanding of gene regulatory networks requires integrating epigenomic datasets from key pathogenic cell types—such as the pituitary gland, granulosa, and theca cells, and potentially, the hypothalamus—across follicular phases to map the spatiotemporal regulation of genes involved in steroidogenesis and folliculogenesis. Despite the hypothalamus’s central role in the HPG axis, regulatory networks mediated by GnRH signaling remain poorly understood. Disruptions in this pathway may explain the involvement of risk genes such as *CNTNAP5*, *ASIC2*, and *CUX2*, potentially linking PCOS to prevalent mental health disorders (3). Incorporating these datasets can enable the development of more inclusive deep-learning models capable of predicting regulatory activity changes beyond enhancer disruptions, offering deeper insights into PCOS pathophysiology.

Additionally, our PCOS subtype classification remains incomplete due to lack of data, leaving some loci unassigned, which may exclude crucial transcription factors and interactions essential for understanding regulatory networks. Lastly, our analysis of causal variants was limited to those occurring within putative enhancers. However, variants can impact gene regulation beyond enhancer activity. Variants located in silencers or insulators may disrupt distal enhancer interactions, as observed with IRX3, emphasizing the need for Hi-C data from pathogenic and affected cell types to resolve target genes not identifiable through eQTL analysis. Lastly, a comprehensive approach should also consider the trans-regulatory effects of risk variants—whether through TFs encoded by susceptibility loci (*PROX1*, *SOX5/8*, *IRF1*) or non-coding RNAs that contribute to epigenomic regulation of gene expression.

### Conclusions

Our results provide valuable insights into molecular mechanisms underlying PCOS etiology. Future *in vitro* and *in vivo* characterization will be essential to validate these predictions, potentially paving the way for novel, symptom-targeted therapies for PCOS patients.

## Methods

### PCOS susceptibility loci

PCOS GWAS summary statistics were obtained from the NHGRI-GWAS catalog. Variants in LD were expanded and clustered into 50 loci based on 100kb proximity. Subtypes identified for 38 GWAS variants (Table S4) were also assigned to their LD variants. The risk allele from GWAS summary statistic served as the alternate allele for GWAS variants, while the minor allele from the 1000Genomes catalog was assumed as risk allele for LD variants. All analyses were conducted using the coordinates and datasets of GRCh38 reference genome.

### Transcription factor binding sites

Transcription factor binding site (TFBS) regions were defined by extending variant sites by 30 bp on each side. TF binding profiles from HOCOMOCO (71) and JASPAR non redundant collection (72) were analyzed using FIMO with default parameters (73). Aside from gain and loss of motifs, changes in motif scores were used to assess affinity differences between reference and alternate alleles. A list of all the TFBSs gained, lost and modulated for SNPs exhibiting significant fold change is provided in Table S9.

### Cell type specific DL models

We used a two phase TREDNet model developed in our lab for cell-type specific enhancer prediction (74). The first phase of the model was pre-trained on 4560 genomic and epigenomic profiles, which included DHS, ATAC-Seq, Histone ChIP-Seq and and TF ChIP-Seq peaks from ENCODE v4 (75). The second phase was fine-tuned to predict cell type specific enhancers using training datasets described below. Chromosomes 8 and 9 were held out for testing, chromosome 6 was used for validation and other autosomal chromosomes were used to build the second phase model. The area under the ROC and PRC curve for each of these models is provided in Figure 2A. The pre-trained phase-one model has been deposited at https://doi.org/10.5281/zenodo.8161621.

Open chromatin (DHS or ATAC-Seq) and H3K27ac profiles for the causal cell-types were downloaded from ENCODE (75) (Table S8). Positive datasets were defined as 2 kb regions centered on DHS or ATAC-Seq peaks overlapping with H3K27ac (or H3K4me1 in fetal brain) peaks of each cell type, excluding coding sequences, promoter proximal regions (<2kb from TSS) and ENCODE blacklisted regions (76). A 10-fold control dataset was generated for each cell-type using randomly sampled 2kb fragments of the genome, excluding the positive dataset of that cell type and blacklisted regions.

Each 2 kb fragment received an enhancer probability score. Active enhancers were predicted at a 10% FPR with a 1:10 positive-to-control ratio. Variant effects were assessed by scoring 2 kb regions centered on each variant for reference and alternate alleles. A significant enhancer activity change was defined as an alternate/reference score ratio >1.2 or <0.8.

### Enrichment analysis of TFBSs

We used command line FIMO (77) to scan vertebrate TF motifs from JASPAR (78) and HOCOMOCO (79) databases along the sequences, applying a p-value threshold of 10^-5^. To identify TFs enriched in the loci of pcosSNVs, we generated a background set of SNVs by extracting all variants from the 1000 Genomes Project within a 50 kb flanking region of each pcosSNV. After excluding the pcosSNVs themselves and removing duplicates, this resulted in a non-redundant background set of approximately 71,000 SNVs. Differential enrichment of TFBSs between the metabolic and reproductive subtypes was assessed using a binomial test, with normalized counts of a TF overlapping variants of one subtype analyzed against the normalized counts of the same TF overlapping variants of the other subtype as the background.

### GWAS trait enrichment

Summary statistics for 25,649 traits were downloaded from the NHGRI-GWAS catalog. Linkage disequilibrium (LD) variants for each GWAS SNP were identified using PLINK (v1.9(80)) with an r² threshold of ≥ 0.8. Traits with at least 1,000 combined GWAS and LD variants were retained for downstream enrichment analysis in reproductive and metabolic SNV categories.

### Data and tools

The H3K27ac peaks for KGN cells and adipocytes were sourced from literature (81,82). The KGN wig file was converted to NarrowPeak format using UCSC BigWig tools (83) and MACS peak calling software (84).

Motif logos were retrieved from HOCOMOCO database (79). Ontology enrichment of pcosSNVs was performed using the Molecular Signatures Database (85). Protein interaction networks and enriched pathways (Figure 3E) were obtained from STRING database (86).

Evolutionary conservation of genomic regions was measured by their extent of overlap with phastCons elements conserved across 30 primates (https://hgdownload.soe.ucsc.edu/goldenPath/hg38/database/ phastConsElements30way.txt.gz).

## Acknowledgements

This work utilized the computational resources of the NIH HPC Biowulf cluster.

## Data availability

Please see the section “Data and tools” and supplementary tables. The deep learning models for eleven cell-types trained in the study are deposited at https://doi.org/10.5281/zenodo.15041688.

## Authors’ contributions

J.S. performed the computational analysis, analyzed the data, and prepared figures and tables. I.O. supervised the study. J.S. and I.O. wrote the manuscript.

