## Supplementary material for "Regulatory risk loci link disrupted androgen response to pathophysiology of Polycystic Ovary Syndrome": Figure S

**This PDF file includes:**

Supplementary figures S1 to S9

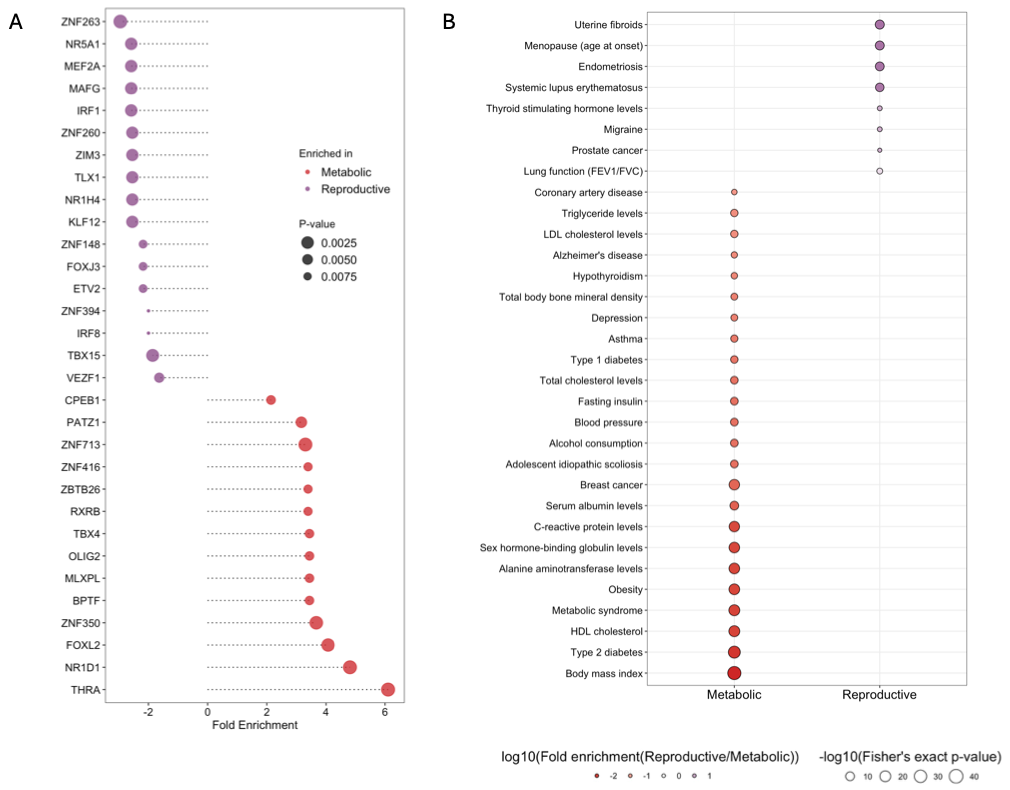

**Figure S1:** (A) Variation in TFBS enrichment between metabolic and reproductive subtypes, (B) GWAS traits enriched in each subtype. All reported terms meet the significance threshold of binomial p-value < 0.01.

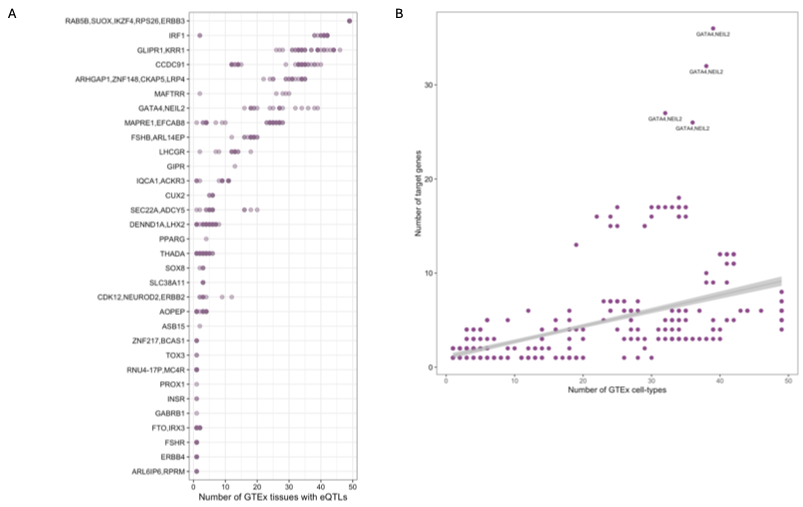

**Figure S2:** (A) Number of eQTLs overlapping with pcosSNVs in risk loci, (B) Correlation between the number of target genes regulated by eQTLs and the number of cell-types in which target gene(s) expression is affected.

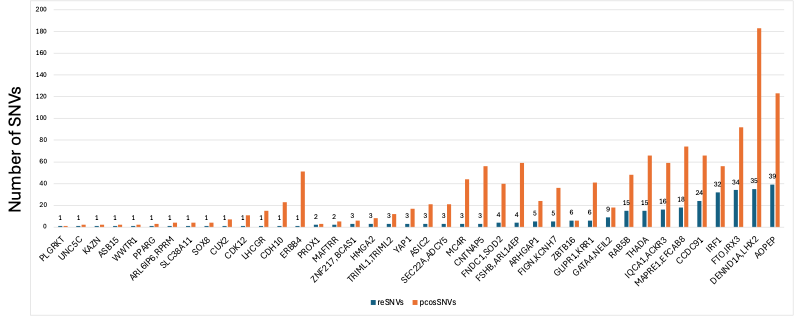

**Figure S3:** A comparison of number of pcosSNVs and reSNVs prioritized by TREDNet.

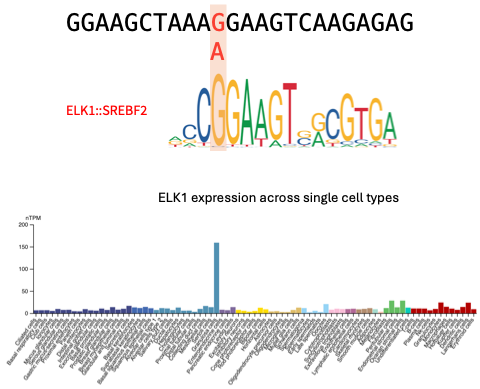

**Figure S4:** The G 🡪 A variant (rs113168128) in ERBB4 locus is predicted to disrupt the binding site of ELK1:SREBF2 motif (Top); Gene expression of ELK1 across single cells obtained from the Human Protein Atlas.

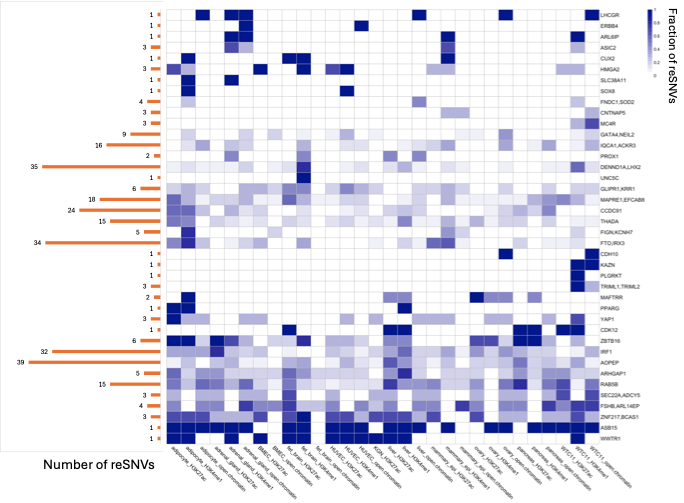

**Figure S5:** Heatmap depicting the overlap of reSNVs in the PCOS risk loci with active regulatory regions of eleven cell-types. Total number of reSNVs predicted by TREDNet in each locus are indicated as bar plot on the left.

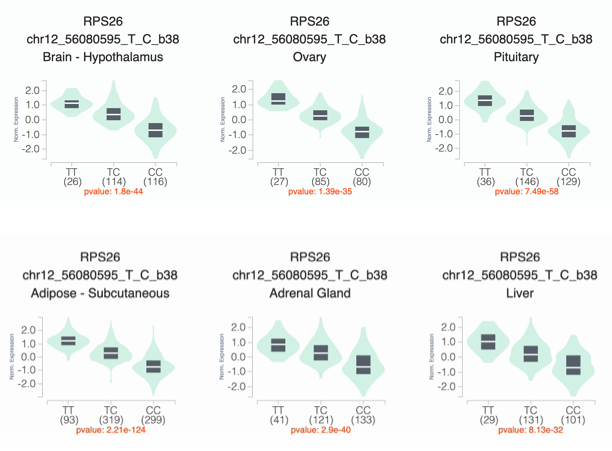

**Figure S6:** Comparison of normalized effect sizes for gene expression of RPS26 across six GTEx tissues impacted by the eVariant rs3741499, a TREDNet-prioritized reSNV.

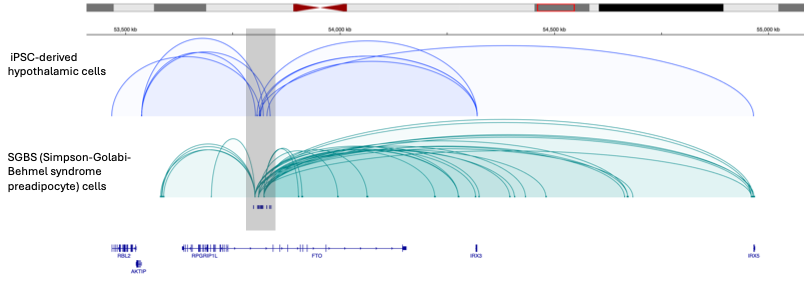

**Figure S7:** Hi-C based interactions between reSNVs of FTO locus (grey bar) with IRX3 and IRX5 in hypothalamic neurons and pre-adipocytes obtained from ref. 62.

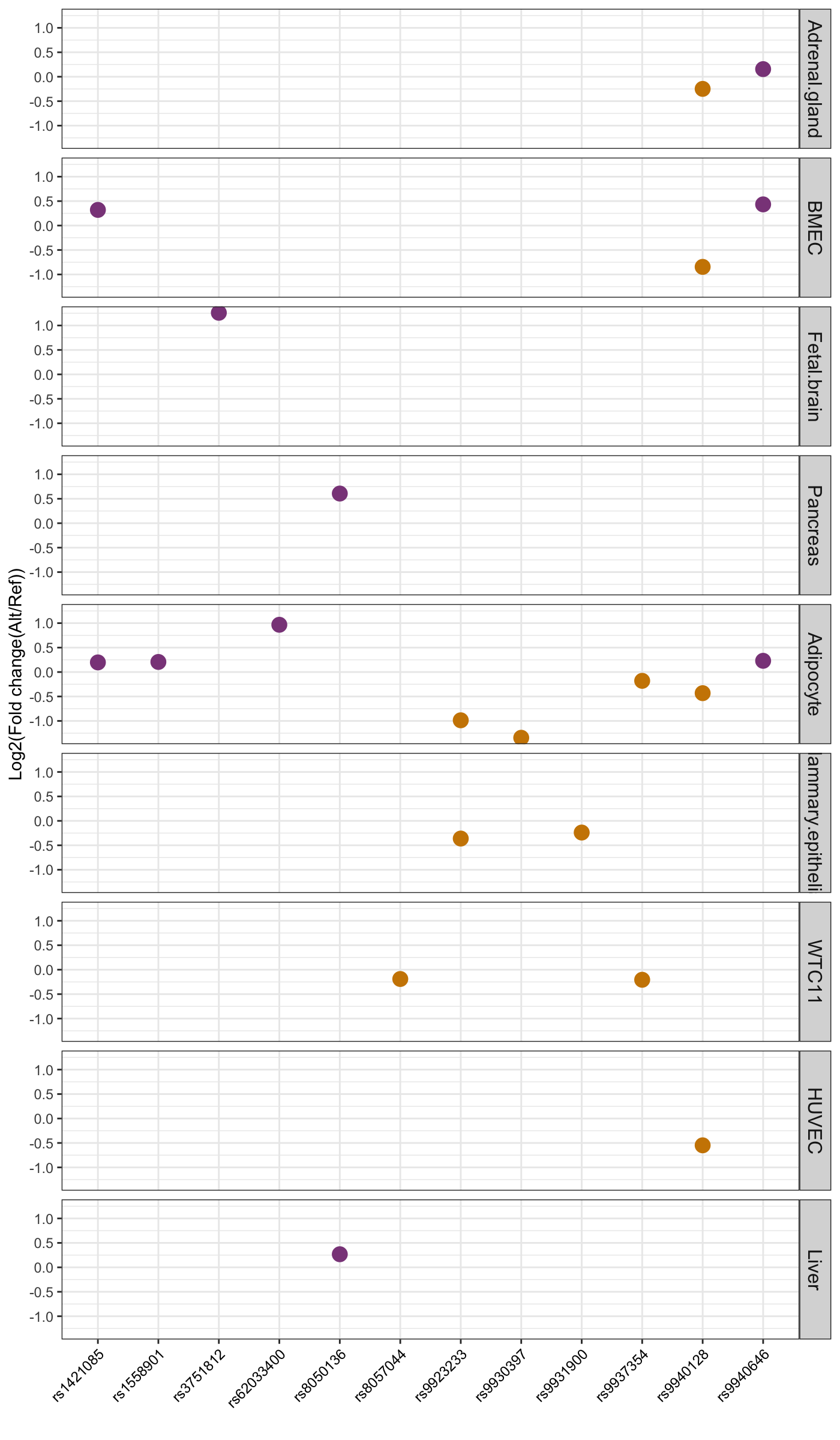

**Figure S8**: Change in enhancer activity between reference and alternate alleles of reSNVs predicted by TREDNet in the FTO locus. reSNVs in this locus were identified in nine cell-types. Purple and orange dots indicate enhancer strengthening and disrupting mutations, respectively.

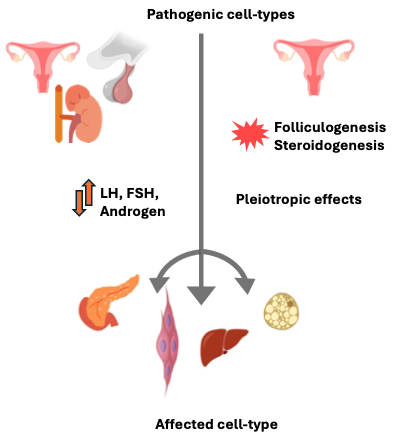

**Figure S9:** Proposed PCOS associated mechanisms linking pathogenic cell-types in disease etiology.
